# Inotropes and mortality in patients with cardiogenic shock: An instrumental variable analysis from the SWEDEHEART registry

**DOI:** 10.1101/2024.05.06.24306966

**Authors:** Petur Petursson, Thorsteinn Gudmundsson, Truls Råmunddal, Oskar Angerås, Araz Rawshani, Moman A. Mohammad, Jonas Persson, Joakim Alfredsson, Robin Hofmann, Tomas Jernberg, Ole Fröbert, David Erlinge, Björn Redfors, Elmir Omerovic

## Abstract

**Background:** The use of inotropic agents in treating cardiogenic shock (CS) remains controversial. We aimed to investigate the effect of treatment with inotropes on 30-day mortality in patients with CS from the SWEDEHEART registry (The Swedish Web-system for Enhancement and Development of Evidence-based care in Heart disease Evaluated According to Recommended Therapies).

**Methods:** We used data from the national SWEDEHEART registry on all patients diagnosed with CS in Sweden between 2000 and 2022. The primary endpoint was 30-day all-cause mortality. We used multilevel Cox proportional-hazards regression with instrumental variable and inverse probability weighting propensity score to adjust for known and unknown confounders. The treatment-preference instrument was the quintile of preference for using inotropes at the treating hospital.

**Results:** In total, 16,214 patients (60.5% men and 39.5% women) were included; 23.5% had diabetes, 10.2% had a previous myocardial infarction (MI), and 13.8% had previous heart failure (HF). The median age was 70 years (interquartile range; 19), and 66.4% were >70. Acute coronary syndrome (ACS) was the cause of CS in 82.9% of patients. Inotropic agents were used in 43.8% of patients, while 56.2% did not receive inotropic agents. There were 7,875 (48.1%) deaths. On average, patients treated with inotropes were two years younger and more likely to have ACS. Patients not treated with inotropes were more likely to have previous MI and previous PCI but less likely to undergo PCI. The number of patients with CS decreased by 12% per year (P_trend_<0.001). There was a considerable variation between hospitals in the preference for using inotropes ranging from 25 to 78% (P<0.001). Inotropes increased by 5% per year (P_trend_<0.001). The unadjusted mortality in CS increased by 2% per calendar year (P_trend_<0.001). The risk of death was higher in patients treated with inotropes [adjusted hazard ratio (HR_adj_) 1.72; 95% confidence interval (CI) 1.26-2.35; P=0.001]. There was a quantitative interaction between inotrope treatment and age and diagnosis (P_interaction_ < 0.001 and P_interaction_ = 0.018, respectively).

**Conclusions:** In this observational study, using inotropes was associated with a higher mortality risk in patients with CS. The increased risk of death was more pronounced in patients younger than 70. The number of patients with CS is decreased, while the use of inotropes and mortality increased in Sweden.

## I. Introduction

Cardiogenic shock (CS) is a severe manifestation of heart failure that can occur due to various cardiac conditions, including acute myocardial infarction, severe valvular disease, and cardiomyopathies^1–4^. Despite advances in managing various cardiac disorders, the CS mortality rate remains high, ranging from 40% to 50%^5–7^. The mortality in CS has remained high over the last few decades. Inotropic agents are commonly administered in CS to improve cardiac output and perfusion^8^. However, their use remains controversial due to concerns about their potential adverse effects, including arrhythmias, myocardial ischemia, and increased mortality^7^. This study aimed to investigate the effect of treatment with inotropes on mortality in patients with CS using data from the Swedish Web-system for Enhancement and Development of Evidence-based care in Heart disease Evaluated According to Recommended Therapies (SWEDEHEART) registry^9^.

## II. Methods

### A. Study Design and Data Source

We conducted an observational study including all patients diagnosed with CS between 2000 and 2022 registered in the SWEDEHEART registry^9^ (Figure 1).

**Figure 1.**
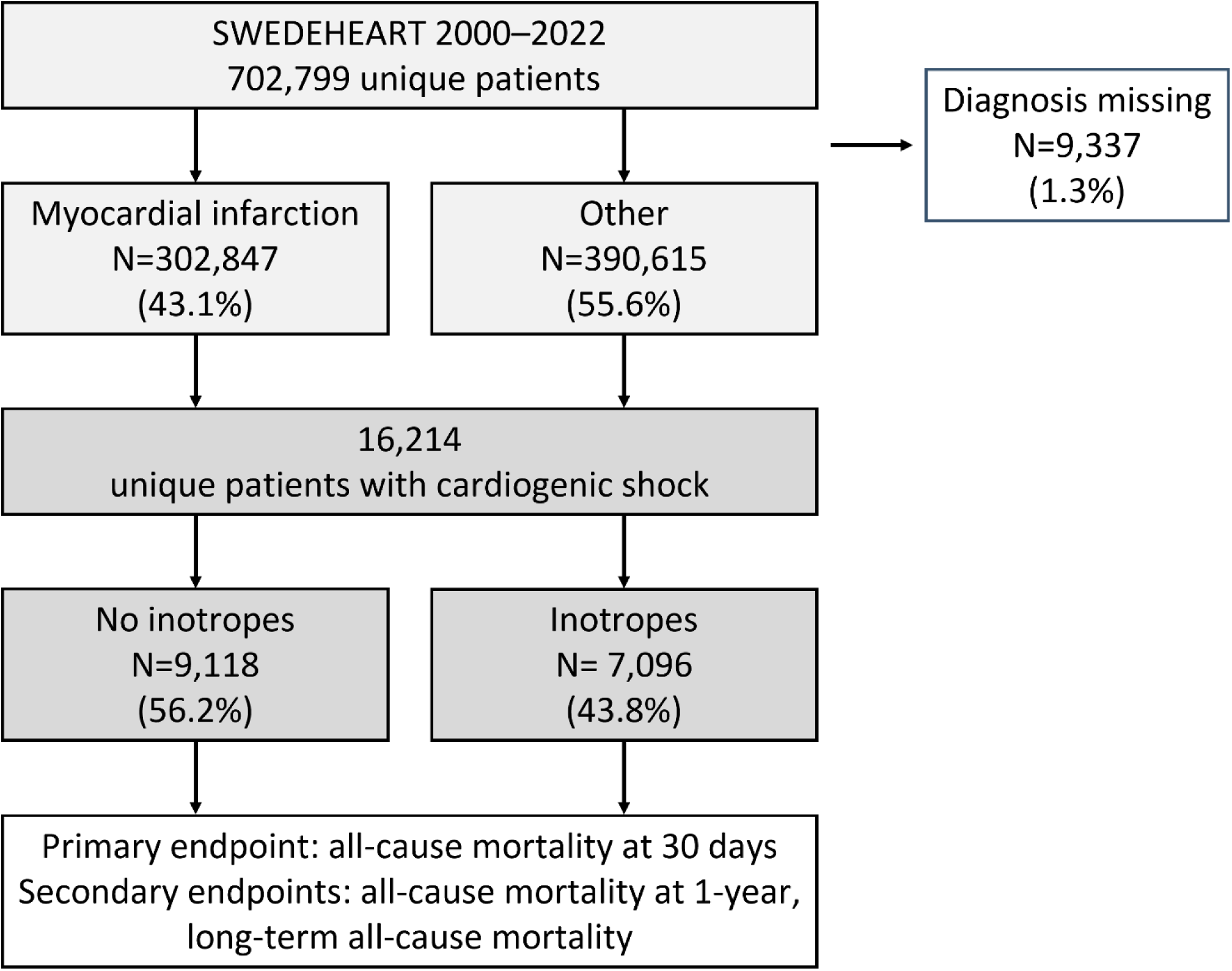
The figure illustrates the patient selection process from the SWEDEHEART registry between 2000-2022. Of 702,799 unique patients, 9,337 (1.3%) had missing diagnoses. Among the remaining patients, 302,847 (43.1%) were diagnosed with myocardial infarction, and 390,615 (55.6%) had other diagnoses (unstable angina, stable angina, observation for chest pain, myocardial infarct complications, heart failure, arrhythmias, valve disease, myocarditis, cardiomyopathies). Among these, 16,214 unique patients were identified with CS. Subsequent treatment categorization revealed that 9,118 patients (56.2%) did not receive inotropic agents, while 7,096 patients (43.8%) were treated with inotropic agents.

### B. Study Population and Variables

We identified patients with CS treated in cardiac care units using the International Classification of Diseases, Tenth Revision (ICD-10) codes I21.0 and I50.1. The primary exposure of interest was inotrope use, defined as the administration of noradrenaline, dobutamine, dopamine, milrinone, or any other inotropic agent during hospitalization for CS. CS was defined as SBP less than 90 mmHg or need of vasopressor therapy to achieve a blood pressure at least 90 mmHg; pulmonary congestion or elevated left-ventricular filling pressures; signs of impaired organ perfusion in a normovolemia or hypervolemia state, with at least one of the following criteria: altered mental status, cold, clammy skin; oliguria; and increased serum lactate. The primary outcome was 30-day all-cause mortality. The registry collects data on demographic characteristics, comorbidities, clinical presentation, hospital characteristics, and treatment modalities.

### C. Statistical Analysis

We used Cox proportional-hazards regression models adjusted for confounders. To apply a conventional adjustment for selection bias attributable to measured confounding, a multivariable Cox proportional hazards regression was performed on the overall cohort using the following covariates: age at admission, sex, smoking status, history of stroke, chronic kidney disease, history of chronic obstructive pulmonary disease, dementia, heart failure, history of myocardial infarction, diabetes mellitus, peripheral arterial disease, cancer, chronic dialysis treatment, hypertension, coronary artery bypass grafting, percutaneous coronary intervention, diagnosis, coronary angiography, and body mass index. We treated hospitals as a random effect to adjust for patient-clustering within the hospitals.

To account for measurable and unmeasured confounders, we performed instrumental variable (IV) analysis based on the two-stage residual inclusion estimate Cox proportional-hazards regression^10,11^. IV analysis is a statistical method for the causal inference of an exposure or treatment on an outcome when potentially confounding factors or unmeasured variables might bias the outcome^12^. It involves IV, a variable that influences the treatment/exposure but is independent of the outcome. Typically, a two-stage regression model is utilized for IV analysis implementation. Initially, a regression model is fitted with the IV as the predictor variable and the treatment/exposure variable as the outcome variable. In the second stage, the predicted values of the treatment/exposure are used as the independent variable in another regression model. The outcome variable is regressed on the predicted values of the treatment/exposure while adjusting for potential confounding variables. In our investigation, the IV utilized was the quintile of preference for administering inotropes at the treating hospitals. We did subgroup analyses to see how age, gender, diabetes, and type of hospital modified the observed effects.

Every analysis adhered to the accepted definition of statistical significance, which is a 2-tailed α =0.05. Version 4.3.0 R (Foundation for Statistical Computing) was used for all statistical calculations and data visualization. The lmtest and ivtools packages were used for 2-stage residual inclusion analysis and IV validity testing. The survival and survminer packages were used for survival analyses, the gtsummary package for creating tables, and the forestploter package for creating forest plots.

Missing values in the dataset were imputed using the missRanger package in R, which utilizes random forest imputation to fill in missing values^13^. The missRanger method has been shown to produce accurate imputations while preserving the data distribution, making it a reliable approach to missing data imputation^13^.

We performed sensitivity analyses to assess our findings’ robustness and the potential impact of misclassification and unmeasured confounding^14,15^. For this purpose, we utilized two R packages: tipr and episensr.

## III. Results

### A. Study Population

We included 16,214 patients with CS in the analysis (Figure 1). The summary of patients’ characteristics is presented in Table 1 and medication at admission is presented in Table 2. Generally, a standardized mean difference (SMD) of 0.15 or lower is considered small. The median age was 70 years (interquartile range; 19), and 66.4% were >70. The most common cause of CS was acute coronary syndrome (ACS) (82.9%). Inotropic agents were used in 43.8% of patients, while 56.2% did not receive inotropic agents. The only variables with SMD > 0.1 between the two groups were age, sex, smoking, diagnosis, angiography, and PCI. On average, patients treated with inotropes were two years younger and more likely to have ACS. Patients not treated with inotropes were less likely to undergo angiography and PCI and were more likely to be smokers. There were no substantial differences in drugs at the time of admission between the two groups. The number of patients with CS decreased by 12% per calendar year (P_trend_<0.001). The use of inotropes increased by 5% per calendar year (P_trend_<0.001, Figure 2A).

**Figure 2.**
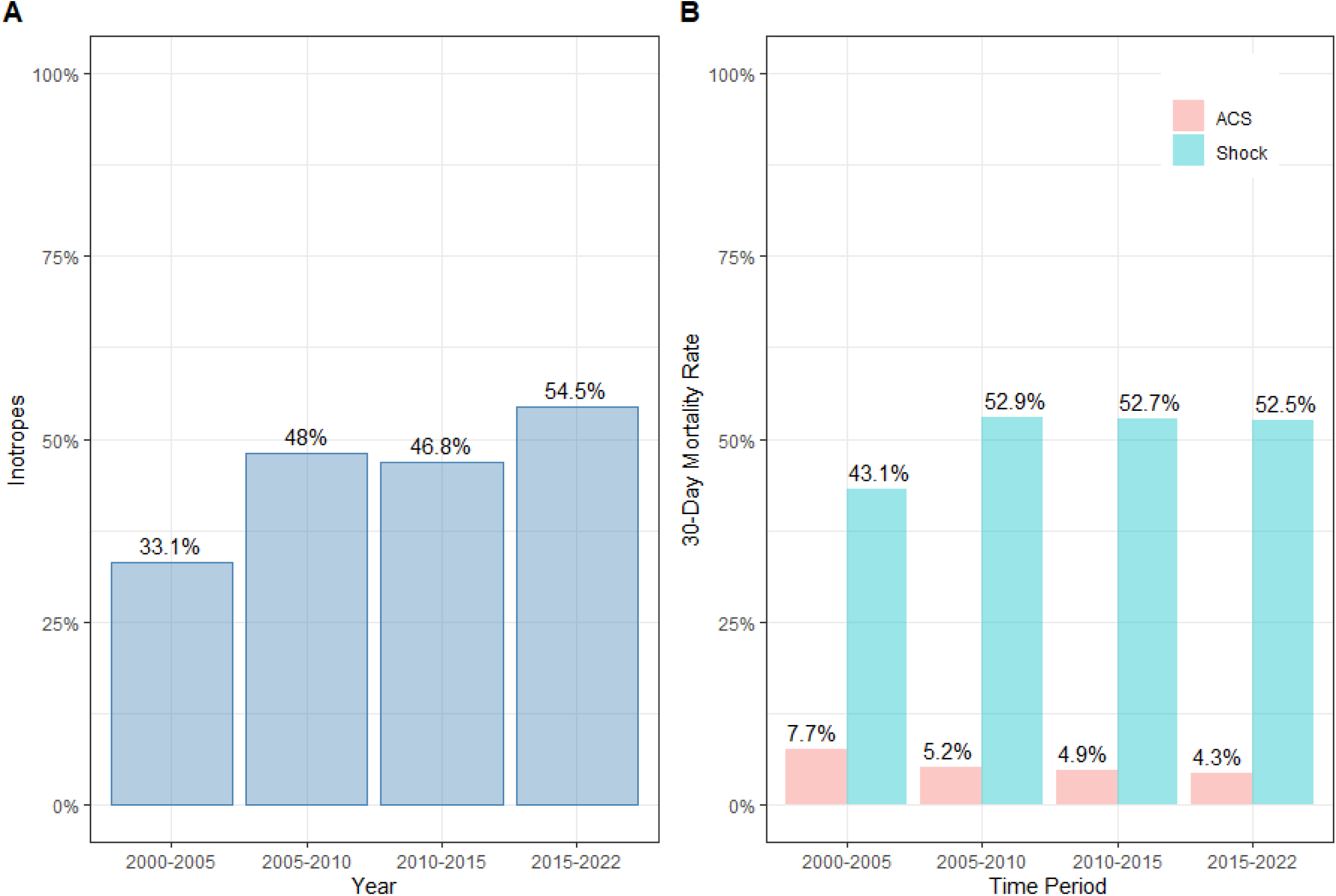
Trends in utilization of inotropes and mortality in Sweden 2000-2022. Panel A shows the average use of inotropes over time. The y-axis shows the proportion of patients who received inotropes, while the x-axis displays the different periods. Inotropes increased over time, with the highest use during the last period. Panel B depicts the 30-day mortality rate over time, stratified by shock status and ACS diagnosis. The y-axis shows the proportion of patients who died within 30 days of hospital admission, while the x-axis displays the different periods. The figure demonstrates that the mortality rate decreased over time in patients with ACS but remained high in patients with shock.

**Table 1.** Patient’s characteristics.

| Variable | N | No inotropes<br>N = 9118<sup>1</sup> | Inotropes<br>N = 7096<sup>1</sup> | Difference<sup>2</sup> | 95% CI<sup>2,3</sup> |
| --- | --- | --- | --- | --- | --- |
| Age | 16,214 | 76 (66, 83) | 74 (65, 80) | 0.14 | 0.11, 0.17 |
| Male sex | 16,214 | 5,315 (58%) | 4,511 (64%) | 0.11 | 0.08, 0.14 |
| Diabetes | 14,982 | 1,670 (19%) | 1,491 (23%) | 0.10 | 0.06, 0.13 |
| Smoking | 15,308 |  |  | 0.16 | 0.13, 0.19 |
| *No* |  | 3,843 (45%) | 2,647 (39%) |  |  |
| *Yes* |  | 2,021 (24%) | 1,522 (23%) |  |  |
| *Previous* |  | 1,691 (20%) | 1,521 (23%) |  |  |
| Stroke | 16,214 | 1,165 (13%) | 814 (11%) | 0.04 | 0.01, 0.07 |
| Renal failure | 16,214 | 328 (3.6%) | 274 (3.9%) | 0.01 | -0.02, 0.04 |
| COPD | 16,214 | 683 (7.5%) | 537 (7.6%) | 0.00 | -0.03, 0.03 |
| Dementia | 16,214 | 80 (0.9%) | 36 (0.5%) | 0.04 | 0.01, 0.08 |
| Heart failure | 16,214 | 1,378 (15%) | 863 (12%) | 0.09 | 0.06, 0.12 |
| Myocardial<br>infarction | 16,214 | 985 (11%) | 675 (9.5%) | 0.04 | 0.01, 0.07 |
| PAD | 16,214 | 596 (6.5%) | 437 (6.2%) | 0.02 | -0.02, 0.05 |
| Cancer | 16,214 | 443 (4.9%) | 291 (4.1%) | 0.04 | 0.01, 0.07 |
| Dialysis | 16,214 | 77 (0.8%) | 57 (0.8%) | 0.00 | -0.03, 0.04 |
| Hypertension | 16,214 | 2,172 (24%) | 1,690 (24%) | 0.00 | -0.03, 0.03 |
| Previous CABG | 16,214 | 285 (3.1%) | 241 (3.4%) | 0.02 | -0.02, 0.05 |
| Previous PCI | 16,214 | 120 (1.3%) | 103 (1.5%) | 0.01 | -0.02, 0.04 |
| Diagnosis | 16,085 |  |  | 0.31 | 0.28, 0.34 |
| MI |  | 6,617 (73%) | 5,971 (85%) |  |  |
| UA |  | 207 (2.3%) | 63 (0.9%) |  |  |
| SA |  | 377 (4.2%) | 161 (2.3%) |  |  |
| HF |  | 515 (5.7%) | 277 (3.9%) |  |  |
| Arrhythmia |  | 532 (5.9%) | 273 (3.9%) |  |  |
| Other |  | 812 (8.9%) | 284 (4.0%) |  |  |
| Angiography | 16,214 | 3,652 (40%) | 4,252 (60%) | 0.41 | 0.37, 0.44 |
| PCI performed | 16,214 | 2,839 (31%) | 3,643 (51%) | 0.42 | 0.39, 0.45 |

<sup>1</sup>n (%); Median (IQR),
<sup>2</sup>Standardized Mean Difference
<sup>3</sup>CI = Confidence Interval
COPD = chronic obstructive lung disease, PAD = peripheral artery disease, CABG = coronary artery by-pass surgery, PCI = percutaneous coronary intervention, MI = myocardial infarction, UA = unstable angina, SA = stable angina

**Table 2.** Medication at admission.

| Variable | N | No inotropes<br>N = 9118<sup>1</sup> | Inotropes<br>N = 7096<sup>1</sup> | Difference<sup>2</sup> | 95% CI<sup>2,3</sup> |
| --- | --- | --- | --- | --- | --- |
| **ACEI** | 16,036 | 1,806 (20%) | 1,380 (20%) | 0.10 | 0.07, 0.13 |
| **ARB** | 9,166 | 595 (13%) | 601 (13%) | 0.06 | 0.02, 0.10 |
| **Anticoagulants** | 16,030 |  |  | 0.09 | 0.06, 0.12 |
| No |  | 8,194 (91%) | 6,301 (90%) |  |  |
| Warfarin |  | 551 (6.1%) | 404 (5.8%) |  |  |
| Dabigatran |  | 10 (0.1%) | 14 (0.2%) |  |  |
| Rivaroxaban |  | 20 (0.2%) | 17 (0.2%) |  |  |
| Apixaban |  | 59 (0.7%) | 75 (1.1%) |  |  |
| Other |  | 5 (<0.1%) | 2 (<0.1%) |  |  |
| **Aspirin** | 16,045 | 2,922 (32%) | 1,991 (28%) | 0.11 | 0.08, 0.14 |
| **Beta blocker** | 16,033 | 2,877 (32%) | 2,164 (31%) | 0.09 | 0.06, 0.12 |

| Variable | N | No inotropes<br>N = 9118<sup>1</sup> | Inotropes<br>N = 7096<sup>1</sup> | Difference<sup>2</sup> | 95% CI<sup>23</sup> |
| --- | --- | --- | --- | --- | --- |
| Ca<sup>+</sup> antagonist | 16,008 | 1,382 (15%) | 1,125 (16%) | 0.10 | 0.06, 0.13 |
| Digitalis | 16,023 | 610 (6.8%) | 327 (4.7%) | 0.11 | 0.08, 0.14 |
| Diuretic | 16,035 | 2,797 (31%) | 1,901 (27%) | 0.12 | 0.09, 0.15 |
| Statin | 16,026 | 1,409 (16%) | 1,233 (18%) | 0.10 | 0.07, 0.13 |
| Ezetimibe | 15,275 | 30 (0.4%) | 33 (0.5%) | 0.08 | 0.04, 0.11 |
| Nitrate | 16,020 | 973 (11%) | 556 (7.9%) | 0.12 | 0.09, 0.15 |

<sup>1</sup>n (%)
<sup>2</sup>Standardized Mean Difference
<sup>3</sup>CI = Confidence Interval
ACE = angiotensin-converting enzyme inhibitor, ARB = angiotensin II receptor blocker

### B. Inotrope Use and Mortality

The unadjusted mortality rate in patients with CS increased by 2% per calendar year (P_trend_<0.001). While 30-day mortality decreased in patients with ACS (P_trend_<0.001), CS mortality increased after 2005 (p<0.001, Figure 2B). There was a considerable variation between hospitals in preference for using inotropes, ranging from 8% to 80% (P_trend_<0.001, Figure 3). The mean follow-up time was 3.8 years (range 0-22 years). Unadjusted 30-day (p<0.001, Figure 4A) and long-term mortality (p=0.00031, Figure 4B) were higher in the inotrope group.

**Figure 3.**
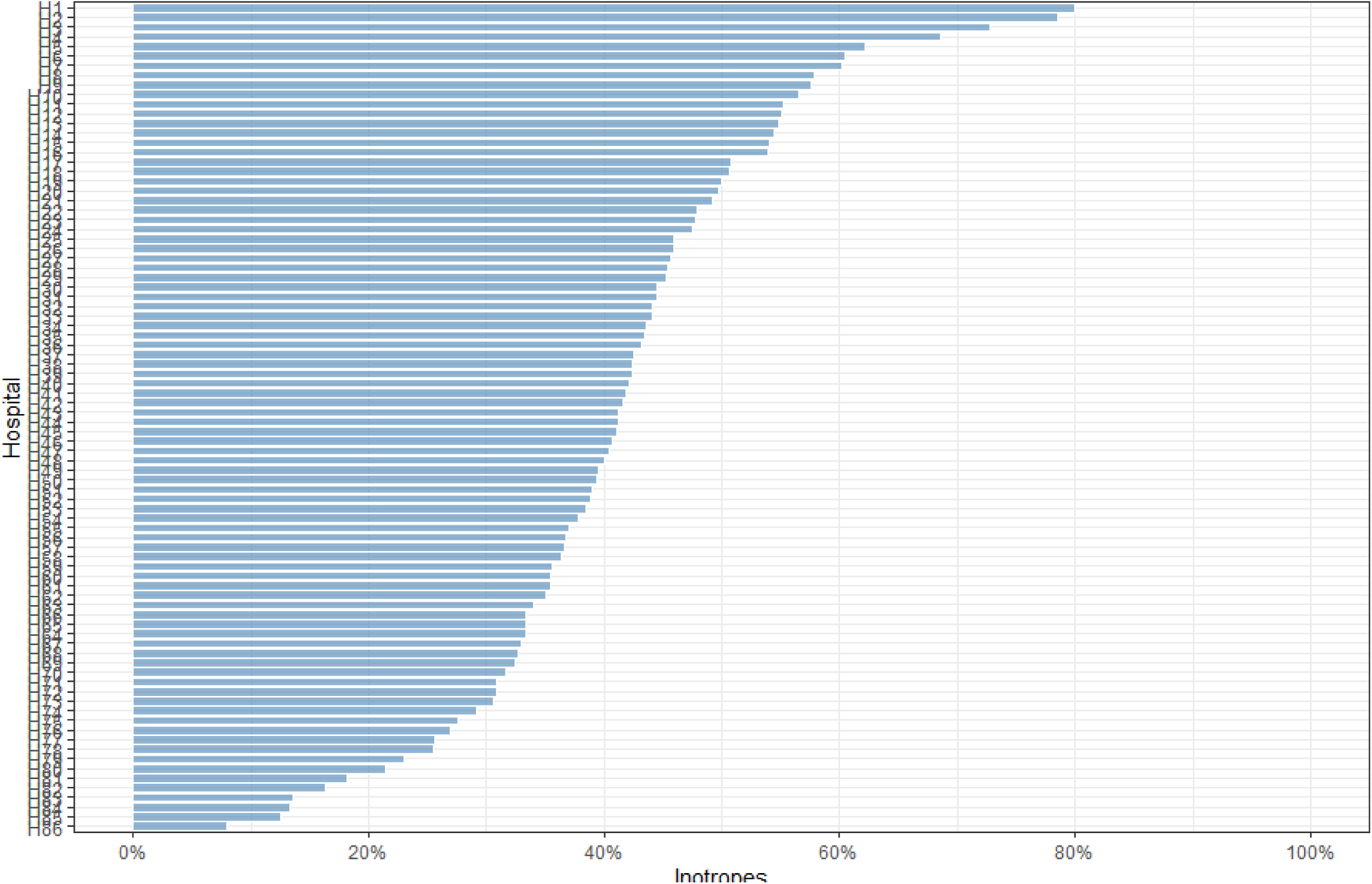
The difference in utilization of inotropes among hospitals in Sweden. The figure illustrates the considerable variation in the preference for using inotropes across different hospitals. The data shows that the use of inotropes varied significantly, ranging from 25% in some hospitals to as high as 78% in others. This considerable variation indicates a lack of standardization in the use of inotropes for managing patients with CS in hospitals.

**Figure 4.**
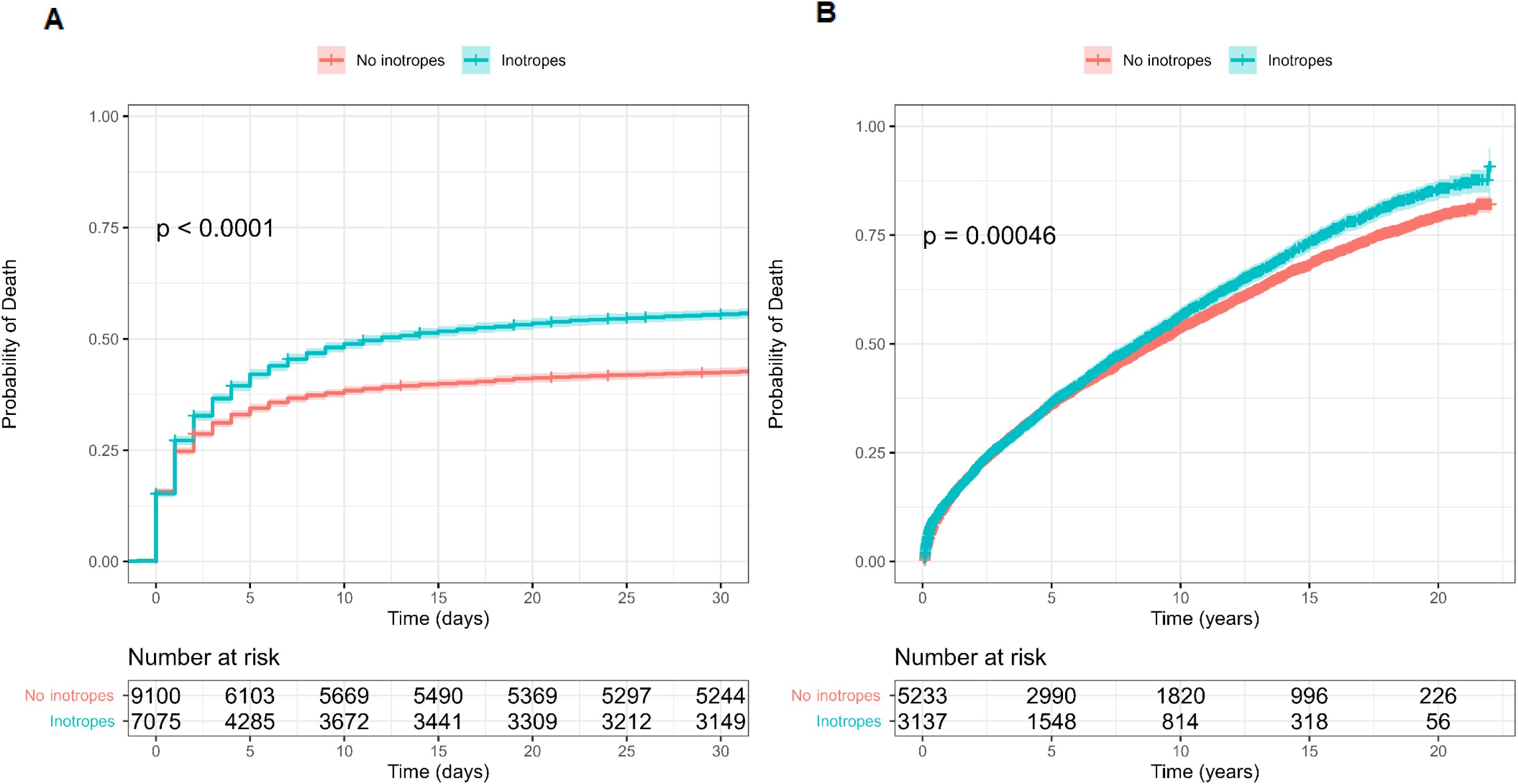
Kaplan-Meier curves for all-cause mortality in patients with CS. Panel A compares 30-day all-cause mortality in patients treated with inotropes (turquoise line) versus those not (orange line). The log-rank test in Panel A demonstrates a significant difference in mortality rates between the two groups (P < 0.001). The risk table below Panel A displays the number of patients at risk every five days for each group. Panel B shows survival after 30 days for the same groups as in Panel A.

#### Instrumental variable analysis

In the first stage of our IV analysis, we identified a significant association between the treatment preference instrument and inotrope use in CS patients (p < 0.001). In the second stage of the analysis, we observed a 72% increase in 30-day mortality linked to inotrope use (Table 3, p < 0.001). Our instrument passed the under-identification test (p < 0.001), suggesting it was relevant. The overidentification test (p = 0.425) did not reject the null hypothesis, suggesting that our instrument was valid. The instrument also passed the weak identification test (Cragg-Donald Wald F statistic= 150, p < 0.001), demonstrating a strong correlation between the instrument and the treatment.

**Table 3.**
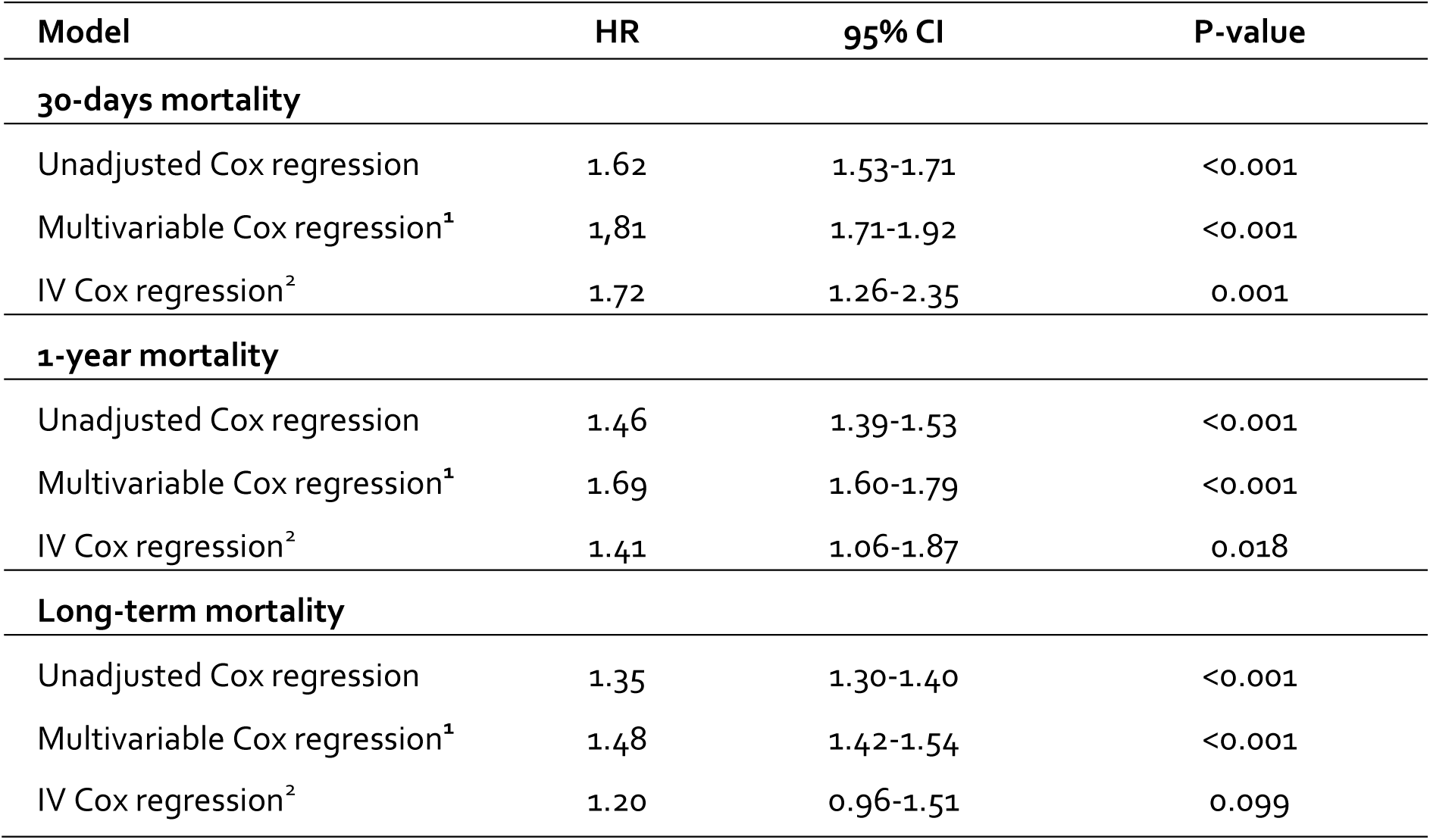
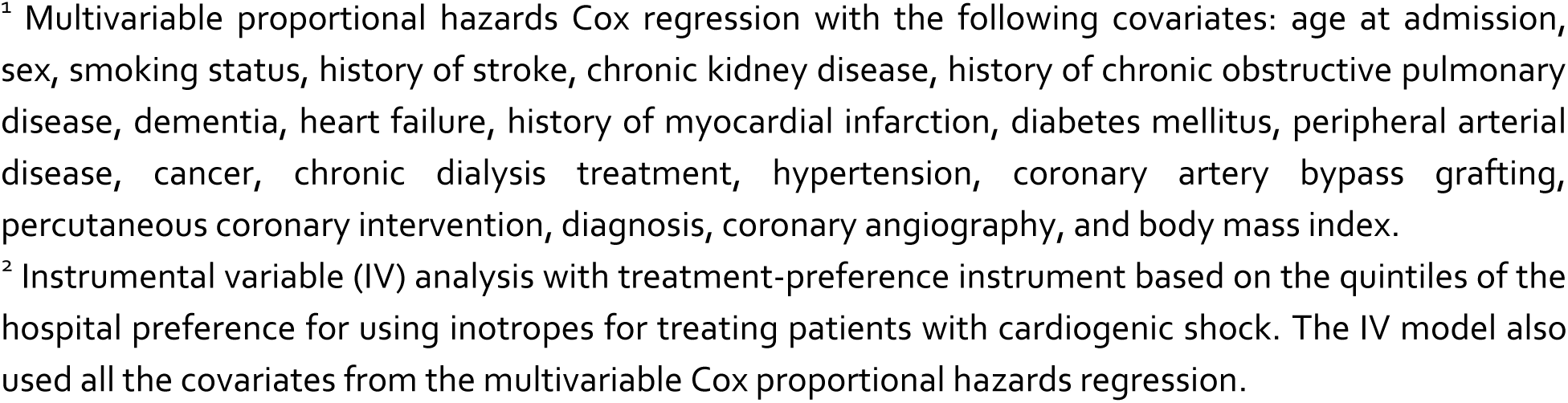
Risk estimates in different statistical models.

| Model | HR | 95% CI | P-value |
| --- | --- | --- | --- |
| **30-days mortality** |  |  |  |
| Unadjusted Cox regression | 1.62 | 1.53-1.71 | <0.001 |
| Multivariable Cox regression<sup>1</sup> | 1.81 | 1.71-1.92 | <0.001 |
| IV Cox regression<sup>2</sup> | 1.72 | 1.26-2.35 | 0.001 |
| **1-year mortality** |  |  |  |
| Unadjusted Cox regression | 1.46 | 1.39-1.53 | <0.001 |
| Multivariable Cox regression<sup>1</sup> | 1.69 | 1.60-1.79 | <0.001 |
| IV Cox regression<sup>2</sup> | 1.41 | 1.06-1.87 | 0.018 |
| **Long-term mortality** |  |  |  |
| Unadjusted Cox regression | 1.35 | 1.30-1.40 | <0.001 |
| Multivariable Cox regression<sup>1</sup> | 1.48 | 1.42-1.54 | <0.001 |
| IV Cox regression<sup>2</sup> | 1.20 | 0.96-1.51 | 0.099 |

<sup>1</sup> Multivariable proportional hazards Cox regression with the following covariates: age at admission, sex, smoking status, history of stroke, chronic kidney disease, history of chronic obstructive pulmonary disease, dementia, heart failure, history of myocardial infarction, diabetes mellitus, peripheral arterial disease, cancer, chronic dialysis treatment, hypertension, coronary artery bypass grafting, percutaneous coronary intervention, diagnosis, coronary angiography, and body mass index.
<sup>2</sup> Instrumental variable (IV) analysis with treatment-preference instrument based on the quintiles of the hospital preference for using inotropes for treating patients with cardiogenic shock. The IV model also used all the covariates from the multivariable Cox proportional hazards regression.

#### Multivariable Cox proportional-hazards regression

The risk of death at 30 days was higher in patients treated with inotropes (adjusted hazard ratio [HR_adj_] 1.81; 95% confidence interval (CI) 1.71-1.92; P<0.001, Figure 1A). The risk of death in the long term was also higher in patients treated with inotropes (HR_adj_ 1.48; 95% CI 1.42-1.54; P<0.001, Figure 1B).

#### Subgroup analysis

We found significant quantitative interaction between treatment with inotropes and age and the cause of CS (Figure 5). Patients >70 years (HR_adj_ 1.54; 95% CI 1.41-1.68; P_interaction_<0.001) and patients in whom CS was caused by ACS (HR_adj_ 1.70; 95% CI 1.58-1.84; P_interaction_=0.018) had a lower risk of death when treated with inotropes. There was no interaction between inotropes and gender, diabetes, or hospital type.

**Figure 5.**
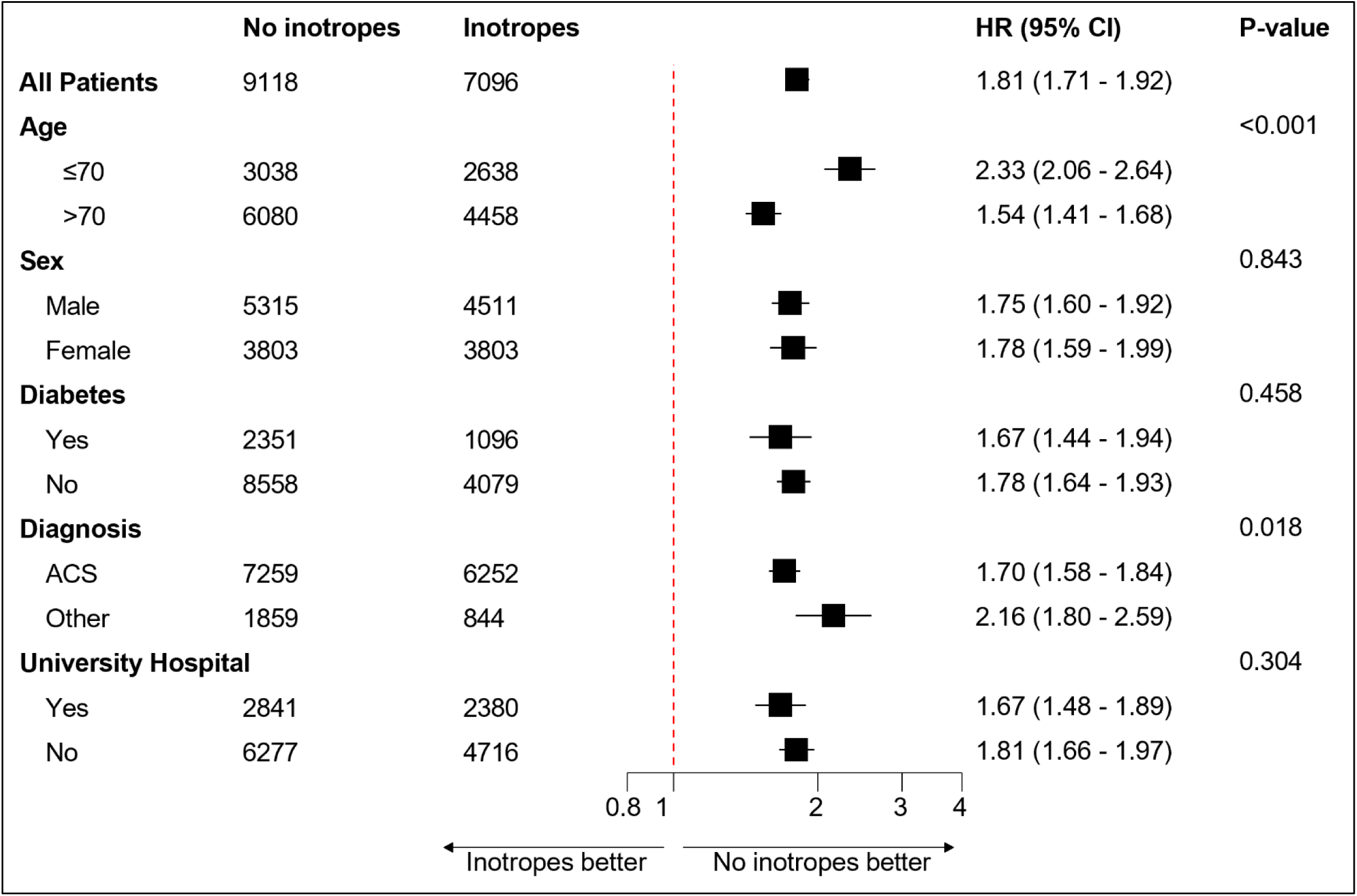
The figure displays a subgroup analysis for age, sex, diabetes, diagnosis, and type of hospital. The data are presented as a forest plot, which summarizes the hazard ratios (HR) and 95% confidence intervals (CI) of the different subgroups on the 30 days mortality. The results of the subgroup analysis indicate that the effect of the inotropes varied across some subgroups. There was evidence for quantitative interaction with a higher risk in younger adults (<70 years) and in patients with cardiogenic shock not caused by ACS.

#### Sensitivity analysis

One or more unmeasured confounders would nullify the observed OR of 1.72 if: 1) the prevalence of the unmeasured confounder in the exposed population was 60%, 2) the estimated prevalence of the unmeasured confounder in the unexposed population was 30%, and 3) an estimated relationship between the unmeasured confounder and the outcome of 9.6 in OR. The misclassification bias for CS diagnosis needed to nullify the OR of 1.72 is 23%. A placebo outcome sensitivity analysis was conducted to validate the instrument using stroke as an unrelated outcome. We found no significant association between inotrope usage and stroke at 30-days (HR 1.0; 95 % CI 0.01-198, P=1.00), at one year (HR 1.0; 95 % CI 0.19-5.10, P=1.00) and long-term (HR 1.0; 95 % CI 0.46-2.06, P=1.00). To evaluate whether treatment-induced selection bias^16^ is affecting the IV risk estimates, we applied inverse probability of selection weights^17^. The estimated risk from the IV model remained substantially unchanged for mortality at 30-days (HR 1.73; 95 % CI 1.26-2.37, P=0.001), at one year (HR 1.38; 95 % CI 1.04-1.84, P=0.027), and long-term (HR 1.19; 95 % CI 0.96-1.49, P= 0.111). These results support the instrument’s validity, implying it is not associated with treatment-induced selection bias.

## IV. Discussion

Our research explored the impact of inotropic therapy on mortality rates, analyzing data from a large cohort of 16,214 patients with CS recorded in the SWEDEHEART registry. We found a strong association between inotrope use and elevated mortality rates in patients with CS. This study represents one of the most comprehensive observational investigations into using inotropes in the context of CS.

The use of inotropic agents in the management of patients with CS has been a subject of controversy for several decades^2,4,18,19^. Inotropes increase myocardial contractility and improve hemodynamics, which makes them an attractive therapeutic option for managing CS^8^. However, some studies have raised concerns about the safety and efficacy of inotropes, suggesting that they may be associated with an increased mortality risk in patients with CS^20–22,7^. Our study highlights the risks associated with inotropic therapy and underscores the need for careful consideration when determining appropriate treatment strategies for CS patients. Our findings are consistent with previous studies reporting that inotropes are associated with an increased risk of mortality in CS^23–26^. One potential explanation for the negative mortality impact is that inotropes increase myocardial oxygen demand, leading to myocardial ischemia and worsening heart failure with consequent hemodynamic instability^1,21,27^. In addition, inotropes may trigger malignant ventricular arrhythmias, further deteriorating the clinical course of patients with CS^6^.

The findings of our study indicate a concerning trend in managing patients with cardiogenic CS. Despite advancements in the treatment for ACS over the past two decades, the mortality rate for patients with CS has remained stagnant at approximately 50%. This lack of improvement in mortality for CS patients contrasts with the significant decrease in mortality for patients with ACS during the same period. One possible explanation for these opposite results is the increased utilization of inotropic therapy. We found a significant relationship between the administration of inotropes and increased short- and long-term mortality rates among CS patients. One could assume that inotrope treatment is used in the most hemodynamically unstable patients. However, all patients in the study were diagnosed with CS and were hemodynamically unstable. The substantial variation between hospitals indicates a lack of standardization in the use of inotropes for managing patients with CS. This difference in preference for inotropes among hospitals has created “natural randomization,” which we could utilize for causal statistical interference with instrumental variable analysis^12^. Several formal tests support the instruments’ validity, and the analyses speak against treatment-induced selection bias^16,17^. Furthermore, the sensitivity analyses demonstrated the IV model’s resistance to residual confounding bias. As a result, our study warns against overusing inotropes in treating CS patients and emphasizes the importance of focusing research on alternative treatments that improve survival.

We found that the higher risk of mortality associated with inotrope use was more pronounced in patients younger than 70 years. Various possible explanations exist for the lower risk in elderly CS patients treated with inotropes. Physicians may be more cautious when treating elderly patients due to the complexity of managing their comorbidities. They may use lower dosages of inotropes, monitor elderly CS patients more closely, and intervene sooner when necessary. More frequent use of other forms of supportive care, such as mechanical ventilation, dialysis, or nutritional support, may also contribute to better outcomes for older adults. However, it is also possible that the lower mortality is due to selection bias, where patients were carefully chosen based on their overall health and likelihood of responding to treatment, making them more likely to survive.

The strength of our paper lies in several aspects, including the large number of death events (~8,000), which provides high statistical power. We utilized instrumental variable analysis, considered the gold standard for statistical causal inference from observational data^12,28,29^. This method allows adjustment for known and unknown confounding factors and mitigates the effects of confounding and selection bias often inherent in observational studies.

### Limitations

While acknowledging the inherent limitations of observational research in establishing causality, our study presents compelling evidence on the relationship between inotrope usage and mortality among patients with CS. One potential criticism of our findings might be the possibility of residual confounding influencing the observed outcomes. However, it is noteworthy that our analysis included adjustments for known predictors of mortality, including comorbidities, which resulted in an increase in risk estimates associated with inotrope use. These results not only strengthen our findings but also match the sharp contrasts seen in baseline characteristics that disadvantage group not treated with inotropes, as shown in Table 1. Patients who did not get inotropes had a much smaller chance of having angiography and PCI, indicating a substantially frailer cohort within the non-inotrope group. This observation lends further support to the argument that the differences in mortality outcomes cannot be solely attributed to unmeasured confounding factors but reflect a genuine disparity in patient outcomes. Moreover, we conduct our study in the Swedish healthcare system, which may limit how well our findings apply to other populations or healthcare systems. The inability to specify the exact type, dosage, and duration of inotrope treatment is acknowledged as a limitation. However, this is mitigated by evidence from recent meta-analyses suggesting a lack of conclusive data favoring one inotrope over others in terms of mortality reduction^30,31^. Another limitation is that we lacked invasive hemodynamic measures in our database, and that some inotropes may have been given for palliative care.

In conclusion, our study does not support using inotropes in managing CS patients. Randomized controlled trials are needed to determine whether inotropes are beneficial, neutral, or harmful in treating CS and to define the optimal type, dosage, and duration of therapy for specific agents. In addition, alternative therapies with a lower risk of adverse outcomes should be explored. To minimize the potential risks associated with inotrope therapy, the cautious use of these agents at the lowest effective dosage and for the shortest duration possible is recommended, or they should be avoided altogether in managing CS.

## Data Availability

Data are not available to the general public due to the Swedish laws and regulations.

